# Chronic Tinnitus is Associated with Aging but not Dementia

**DOI:** 10.1101/2024.07.30.24311207

**Authors:** Lisa Reisinger, Nathan Weisz

## Abstract

**Background:** Aging is related to deterioration of bodily and neural functions, leading to various disorders and symptoms, including the development of dementia, hearing loss, or tinnitus. Understanding how these phenomena are intertwined and how aging affects those is crucial for prevention and the future development of interventions.

**Methods:** We utilized the UK Biobank which includes a total of 502,382 participants between 40 and 70 years old. We used logistic regression models and cox proportional hazard models and compared hazard ratios.

**Results:** The odds of reporting tinnitus in the older age group (i.e., older than 58 years) were increased by 43.3% and a one decibel increase in the SRT enhanced the odds for tinnitus by 13.5%. For our second analysis regarding hearing loss, the risk of dementia increased by 9.2% with an increase by one decibel in the SRT score. In terms of aging, each additional year increased the risk by 19.2%. Tinnitus alone showed a significant influence with a hazard ratio of 52.1%, however, when adding hearing loss, age and various covariates, the effect vanished.

**Conclusion:** Findings confirm that tinnitus is indeed related to aging, but presumably independent of the aging processes accompanying the development of dementia. This highlights the urge to further investigate the impact of aging on neural processes that are relevant for alterations in the auditory systems (e.g., leading to the development of tinnitus or hearing loss) as well as for increased vulnerability in terms of neurodegenerative diseases.

**Key Points:** 

**Question:** Aging and hearing loss have been linked to dementia and tinnitus respectively. But is there a direct influence of tinnitus on dementia risk?

**Findings:** In this case-control study, data derived from the UK Biobank was used to first replicate previous findings establishing aging as a risk factor for tinnitus and hearing loss as a risk factor for dementia. Tinnitus was not found to increase the risk of dementia.

**Meaning:** Aging is related to tinnitus, however, since tinnitus does not influence the risk of dementia, we conclude that the aging processes that determine tinnitus are independent of neural processes facilitating the development of dementia.

## Introduction

Tinnitus is an auditory phantom perception affecting 10-15% of the population and occurs with severe distress in 1-3% including comorbidities such as insomnia, anxiety or depression [1-3]. This circumstance has a high financial impact on society and healthcare systems, including costs for potential treatments or decreased functionality at work, which can lead to early retirements [4]. The financial burden of tinnitus is further bolstered by its heterogeneity which increases the difficulty in developing effective methods for both objective diagnosis and reliable clinical interventions [1]. To date, the mechanisms and neural causes of tinnitus are still not fully understood, impeding both scientific and medical advances in this field and leading to inconclusive results and treatment approaches [5].

It is a wide consensus that to understand tinnitus, the crucial – likely eliciting – role of hearing damage needs to be taken into account. Hearing loss has been diagnosed in 75-80% of tinnitus patients [6], however, these figures likely underestimate the actual relationship, as the diagnosis normally relies on the results of pure-tone audiometry, which do not identify every occurrence of hearing damage [7]. This so-called hidden hearing loss, namely a normal audiogram despite existing hearing impairments, has been additionally widely reported in tinnitus [8,9]. On a conceptual level, the lack of tinnitus in individuals with measurable hearing damage poses greater issues, pointing to additional factors that influence an individual’s risk to develop tinnitus.

Recently, we have argued that aging, or rather some yet-to-be-identified aspect of biological aging, could be a relevant risk factor. This idea is based on the observation that not only does hearing loss increase with age, ranging from a prevalence of 15% up to 60% in older people [10]; but the same tendency is shown in tinnitus with an increased prevalence of 24% in elderly [3,11]. In a recent study, investigating this relationship between hearing loss, tinnitus, and aging [12], we were able to establish aging per se as a tinnitus risk factor. This indicates yet unknown aging-related processes in the brain that facilitate developing tinnitus.

Aging is not only relevant in the context of increased hearing loss and tinnitus but is associated with a great variety of diseases that occur more frequently when we grow older. Dementia as a decline in cognitive abilities affects more than 55 million people with an estimated increase up to 139 million in 2050 [13]. With this continuously increasing prevalence and the major impairments in quality of life coming along with the disease, efficient treatments are urgently needed. However, similar to tinnitus, research is still trying to understand underlying causes and mechanisms as well as potential risk factors. Apart from medical conditions such as diabetes or hypertension [14], aging was established as the most important risk factor as well [15]. Leading back to hearing loss and tinnitus, which are also related to aging, Lin et al. [16] explicitly investigated hearing loss as a risk factor for dementia. Controlling for various confounding variables, hearing loss was found to significantly affect the risk of dementia, which was further supported in recent works (see [17]). In line with the findings reported by Lin et al. [16], age-related hearing loss was reported to affect the risk of dementia [18,19]. If tinnitus is a “symptom” an abnormal brain aging process, then its presence could potentially mark an increased risk to develop dementia. Addressing this question is the main goal of the present study.

Some previous studies suggested that tinnitus could play a role in increased dementia risk. For example, bothersome tinnitus was associated with cognitive impairments [20], presumably mostly because of increased stress and difficulties concentrating that characterizes severe tinnitus [19,21]. In addition, a recent retrospective study demonstrated that, after adjusting for hearing loss, tinnitus increased the risk of early-onset dementia by 63% [22]. The retrospective approach of this study, however, does not allow conclusions regarding causality. As cognitive decline - and further dementia - is linked to a reduction in gray matter [23], Koops and colleagues [24] showed that both hearing loss and age contribute to a decrease in gray matter. Interestingly, however, they additionally demonstrated that this decrease was not observable in patients with tinnitus. Thus, while the influence of hearing loss and aging on tinnitus and dementia are clear, it is unclear whether tinnitus itself increases dementia risk.

In this study, we utilized data from the UK Biobank to analyze the relationships between tinnitus, dementia, hearing loss, and aging. The UK Biobank offers significant advantages due to its large, representative sample and the availability of longitudinal data through access to medical health records, which allows for the assessment of dementia diagnoses over an extended follow-up period. To deepen our understanding of the interplay between these variables, we initially aimed to replicate previous findings, specifically those of Lin et al. [16] and our own earlier work [12]. Using the UK Biobank data, we confirmed our earlier finding that aging is an independent risk factor for tinnitus, irrespective of hearing loss [12]. Additionally, consistent with Lin et al.’s findings, we observed that baseline hearing loss was associated with an increased risk of dementia during the follow-up period. Most importantly however, we found no evidence to support the notion that tinnitus increases the risk of developing dementia.

## Methods

## Participants and experimental procedure

This case-control study has been conducted using data from the UK Biobank (www.ukbiobank.ac.uk), a major biomedical database comprising a total of 502,382 participants which provided questionnaire data, physical tests, genetic information, and imaging data. This information was obtained in four collection periods, allowing for longitudinal analyses as well. This project was accepted under the project ID 90485 and all participants provided written informed consent. For the following analyses, data from the first period and medical health records were used.

For the first analysis, we aimed to replicate the findings of Reisinger et al. [12] to investigate the influence of aging on the risk of developing tinnitus. 122,268 participants between 40 and 70 years (55% female, age mean=56.6 years, SD=8.2 years) were included in the analysis, as information regarding age and hearing status were complete. Additionally, these participants had answered the questionnaire regarding present tinnitus either with “No, never” or “Yes, now most or all of the time”. This selection allowed us to target chronic tinnitus, rather than acute or occasional, to obtain the greatest dissimilarity between participants with and without tinnitus. Hearing loss was assessed using speech-reception thresholds (SRTs), which is a hearing test measuring hearing thresholds at different signal-to-noise ratios. For that, participants were tested in how well they perceived three spoken numbers played with a rushing noise in the background. The final result was defined as the signal-to-noise ratio (in decibel, dB) that allowed the participants to understand half of the presented speech correctly. For the analyses, age was split by the median to generate a younger age group (mean=49.4 years, SD=5.1 years) and an older age group (mean=63.3 years, SD=3.2 years). Sex was added as a covariate. Demographic data is displayed in the Supplementary.

For the second analysis, we aimed to replicate and extend the findings of Lin et al. [16] in order to investigate the influence of hearing loss, and additionally tinnitus, on the risk of developing dementia. 121,045 participants (55% female, age mean=56.6 years, SD=8.2 years) were included in the analysis with completed questionnaires regarding age, hearing status, and tinnitus occurrence (“No, never” or “Yes, now most or all of the time”). As in Lin et al. [16], we included sex, ethnicity, smoking, education, diabetes, and hypertension as covariates. Participants who answered “Do not know” or “Prefer not to answer” in at least one of these variables were excluded from the analyses. Out of these 121,045 participants, 1289 individuals were diagnosed with all cause dementia at least three years after measurements. We aimed to avoid reverse causation in our analyses, hence we excluded participants diagnosed with dementia at the time of measurements and up to three years later. Demographic data is displayed in the Supplementary.

## Statistical Analysis

For the first analysis, we utilized logistic regression models (family=binomial) implemented in R [25]. We aimed to predict tinnitus based on age and hearing status averaged over both ears, adding sex as a covariate. Age was included as a categorical variable by median-splitting the data into two groups (median=58 years). Regression coefficients were transformed into odds ratios to increase interpretability.

For the second analysis, we used the same approach as Lin et al. [16], namely Cox proportional hazards models. In the first, simple model, we targeted the influence of hearing status on the dementia risk as a replication of the results of Lin et al. [16]. We included the covariates sex, usage of hearing aids, hypertension, diabetes, smoking, ethnic, and education as it was implemented by Lin et al. [16]. For a more comprehensive analysis of the risk factors, we used a second, simple model targeting age as a potential risk factor and including the same covariates.

In order to extend the previous findings by Lin et al. [16], we aimed to specifically investigate the influence of tinnitus on the risk of developing dementia. First, we calculated the model with tinnitus as sole predictor. In the next step, we again added the previously described covariates to the model. In a last, combined model, we included all variables mentioned above to obtain a comprehensive overview of the dementia risk factors.

## Results

In the first step, we were able to replicate our results reported in Reisinger et al. [12] and extracted aging as a significant risk factor for tinnitus (*b*=-0.567, *p*<.001). Hearing status (*b*=0.126, *p*<.001) and sex (*b*=0.621, *p*<.001) also were found to be significant predictors for tinnitus. Additional interaction terms did not reveal any significant relations. This result is in line with our previous work; however, sex was an additional risk factor in this sample. The odds of developing tinnitus in the older age group (i.e., older than 58 years) were increased by 43.3% and a one decibel increase in the SRTs enhanced the odds for tinnitus by 13.5%. For sex, men had increased odds by 86.1% to develop tinnitus. In Figure 1A, the influence of age on the risk of developing tinnitus is further displayed. Independent of the hearing status, the tinnitus risk was overall higher in the older age group, bolstering the effect of aging on tinnitus.

**Fig. 1:**
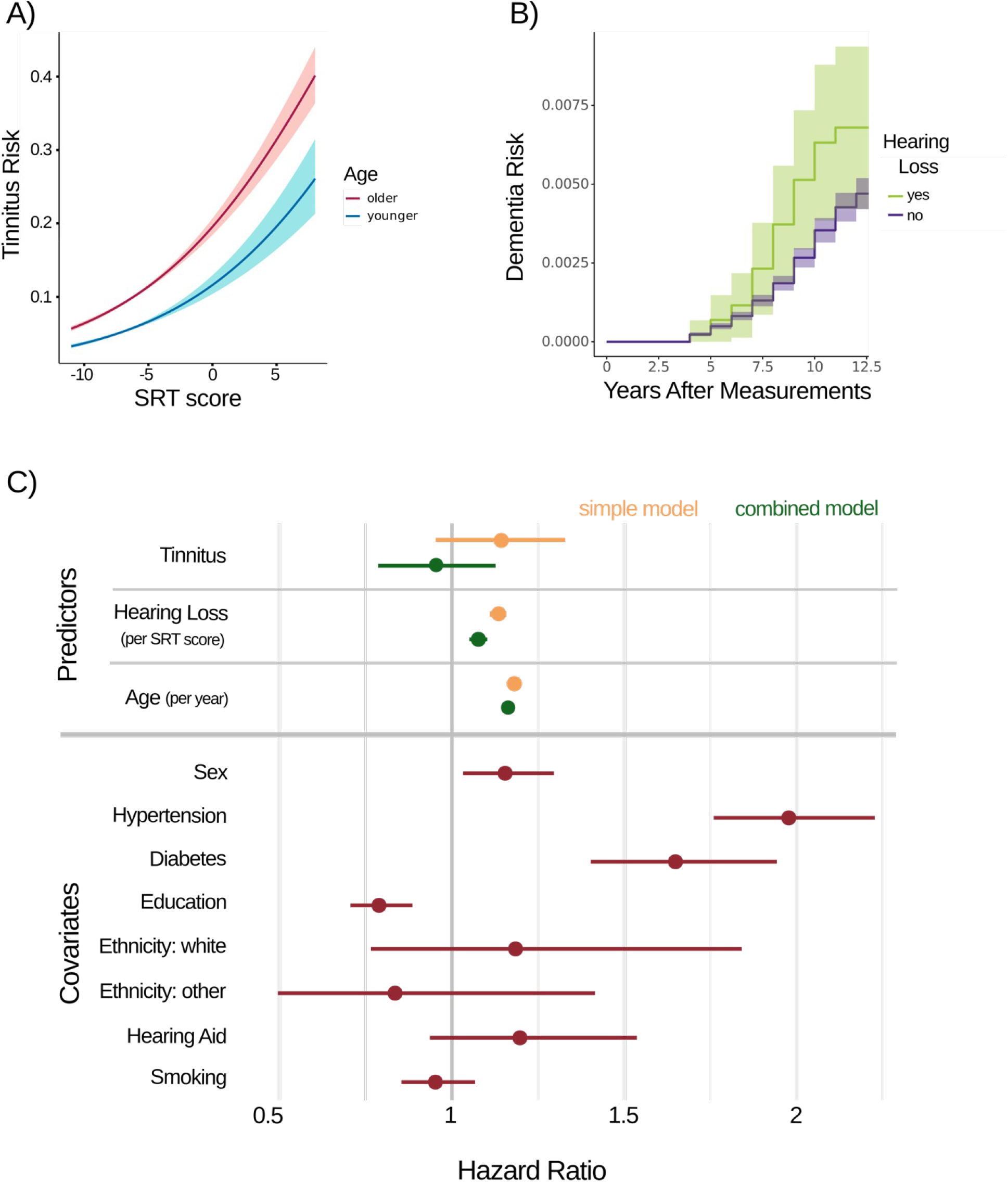
Logistic regression model, Cox proportional hazards model, and hazard ratios. **A)** Logistic regression model. Age characterization was based on the median age of 58 years. The results showed an enhanced probability for tinnitus in the older age group compared to younger people over mean speech-reception-threshold (SRT) scores (in dB). **B)** Cox proportional hazards model. The development of dementia risk after measurements is depicted for individuals with and without hearing loss separately and individuals with hearing problems show a greater risk. Participants with a dementia diagnosis up to three years after measurement were excluded to avoid reverse causation. **C)** Hazard ratios. Hazard ratios for the predictors are displayed for both the simple (i.e. including solely one predictor) and the combined model (i.e. including all three predictors). For the covariates, hazard ratios of the combined model are shown.

In the second analysis, in the first step we were able to replicate the findings of Lin et al. [16] by demonstrating an increased risk of developing dementia by 14.0% (95% CI[1.116, 1.164], p<.001) as hearing ability worsens (i.e., a one decibel increase in SRT scores). Figure 1B demonstrates the increase in dementia risk over years after measurements separately for people with better hearing status and people with worse hearing based on the SRT values (results greater than 0 were considered as hearing difficulties). Further, hazard ratios (HR) indicated that with each additional year, the risk of all-cause dementia increased by 17.3% (95% CI[1.158, 1.187], p<.001).

As a main question of this work, we wanted to investigate the influence of tinnitus on the dementia risk, since tinnitus – as dementia - is highly influenced by aging and hearing loss. In a first model, we aimed to investigate the risk of developing dementia solely based on the occurrence of tinnitus, which showed a significant influence with a hazard ratio of 52.1% (95% CI[1.280, 1.807], p<.001). However, adding the aforementioned covariates to this model, the effect of tinnitus vanished (HR=1.138, 95% CI[0.952, 1.361], p=.155).

In the last step, we implemented a combined model, including tinnitus, hearing status, age, and the covariates. The effects for higher hearing scores (HR=1.078, 95% CI[1.052, 1.104], p<.001) and higher age (HR=1.167, 95% CI[1.152, 1.181], p<.001) increasing the risk of all-cause dementia remained robust in this analysis. Among the covariates, men had a higher risk to develop dementia (HR=1.158, 95% CI[1.034, 1.297], p=.011) and diseases like hypertension (HR=1.981, 95% CI[1.761, 2.229], p<.001) and diabetes (HR=1.652, 95% CI[1.404, 1.945], p<.001) further increased the dementia risk. Better education was found to be a protective factor (HR=0.792, 95% CI[0.707, 0.887], p<.001). Ethnic background (black=1; white: HR=1.188, 95% CI[0.766, 1.843], p=.441; other: HR=0.839, 95% CI[0.497, 1.417], p=.511), smoking (HR=0.956, 95% CI[0.855, 1.069], p=.428), and the usage of hearing aids (HR=1.201, 95% CI[0.938, 1.538], p=.147) did not significantly influence the risk of all-cause dementia. Tinnitus, again, did not influence the dementia risk (HR=0.958, 95% CI[0.803, 1.144], p=.635).

## Discussion

In this work, we aimed both to replicate previous findings regarding the interplay between aging, dementia, tinnitus, and hearing loss, and extend them by novel outcomes as the question arose whether tinnitus has an influence on the risk of developing dementia. Using the UK Biobank, we were able to identify aging as a risk factor for tinnitus, which was independent of hearing loss as the commonly reported main risk factor for tinnitus [26,27]. This effect replicates our previous findings in Reisinger et al. [12], in which we already demonstrated the influence of aging on the tinnitus risk in two independent samples. Using the large amount of data provided by the UK Biobank further bolsters the validity of our effect and highlights the importance to consider aging processes when analyzing tinnitus. Notably, different from our previous results, we found sex as well to be a significant predictor for tinnitus. This is in line with the literature, indicating in general higher tinnitus prevalences in men [28].

Aging as an independent risk factor for tinnitus has now been shown robustly in three different samples (see [12]). Using the UK Biobank for this replication further strengthens the effect in two regards: 1) The assessed sample is older than in our previous work with a median age of 58 years compared to 34 years in the representative sample of our first study (data from the National Health and Nutrition Examination Survey [29]). Hence, the influence of aging has been shown over various age groups. 2) The three analyzed samples utilized three different approaches to assess the hearing status of the participants (online hearing assessment vs. pure-tone audiometry vs. speech-reception thresholds). Despite these variations in the hearing tests, aging was reliably determined as a risk factor independent of the hearing status which supports generalizability of our effect. These differences between the samples and the successful replication using the UK Biobank highlights the influence of aging on the risk of developing tinnitus and calls for urgent research in regard to the underlying aging processes that determine this effect.

The implication that advanced or accelerated aging processes could predispose tinnitus further motivated the question whether tinnitus predicts dementia as an age-related disease [30]. Hearing loss was already associated with an increased dementia risk [16,18,19], and we were able to replicate these findings in our work as well. As for tinnitus, the influence on dementia was significant when including solely tinnitus to the model but vanished when including aging and several covariates. Therefore, our results indicate that aging processes likely separately contribute to increased dementia and tinnitus risk, with the latter not posing an additional risk factor for the development of dementia.

## Limitations and Future Directions

Overall, this work demonstrates the high impact of age on the auditory system as well as on neurodegenerative diseases like dementia. However, conclusions regarding tinnitus are limited since various confounding variables such as age at tinnitus onset or the duration of the auditory phantom perception were not provided by the UK Biobank. Another difficulty to consider when analyzing tinnitus and hearing loss is the occurrence of hidden hearing loss in some individuals [7-9]. This hearing impairment with a normal audiogram is widely undetected and cannot be easily measured objectively. Furthermore, the relationship between hearing loss and dementia could also be influenced by additional variables like social isolation [31]. An active and social lifestyle was proposed to be a protective factor against dementia [32], whereas hearing loss has been widely associated with social isolation as communication grows difficult [33]. Therefore, it is hypothesized that social isolation could be a driving factor in the relationship between dementia and hearing loss and should be taken into account as well [31].

As we reported aging to be of great importance for tinnitus, hearing loss, and dementia, as well as for their complex relationship, we emphasize the necessity to focus future research on aging processes - especially on a neural level. Age-related changes in the brain have been widely reported, also in the context of hearing loss [34-37]. The exact mechanisms remain, however, unclear and it is necessary to unravel causal relationships for a deeper understanding of these processes. For this, novel brain age models and algorithms are crucial to gain new insights into neural processes [38,39]. Moreover, this can further contribute to a deeper understanding of tinnitus. As we have now determined aging as a risk factor, it is still not apparent what exactly increases the risk of tinnitus in an aging brain. Novel findings in this direction can then lead to a better knowledge of this disease and the development of more efficient treatments. Beyond that, preventative approaches could in the first place ensure a healthy aging brain, leading to an overall decreased risk of tinnitus, hearing loss, and dementia.

## Conclusion

The potential influence of aging and hearing loss on dementia and tinnitus has been well-established independently. However, whether these associations suggest a connection between tinnitus and dementia has not been understood until now. We were able to replicate both the effect of aging on tinnitus as well as the effect of hearing loss on dementia. Tinnitus was, however, not established as a risk factor for dementia. Hence, we conclude that tinnitus is indeed related to aging, but presumably independent of aging processes driving the development of dementia. This highlights the urge to further investigate the impact of aging on neural processes that are relevant for alterations in the auditory systems (e.g., leading to the development of tinnitus or hearing loss) as well as for increased vulnerability in terms of neurodegenerative diseases.

## Data Availability

Data was derived from the UK Biobank and cannot be shared by the authors, since the UK Biobank restricts the data access to those researchers with an approved research proposal.

## Supplementary

*Demographic summary of the variables used for the first analysis, split into a tinnitus group and a non-tinnitus group (N=122,268)*.

|  | Tinnitus (n=9520) | No Tinnitus (n=112748) |
| --- | --- | --- |
| SRT value | mean=-6.07 dB (sd=2.23) | mean=-6.68 dB (sd=1.70) |
| Age | mean= 59.5 years (sd=7.23) | mean= 56.4 years (sd=8.18) |
| Age Group | 3130 younger<br>6390 older | 55514 younger<br>57234 older |
| Sex | 5730 male<br>3790 female | 49323 male<br>63425 female |

*Demographic summary of the variables used for the second analysis, split into a dementia group and a non-dementia group (N=121,045)*.

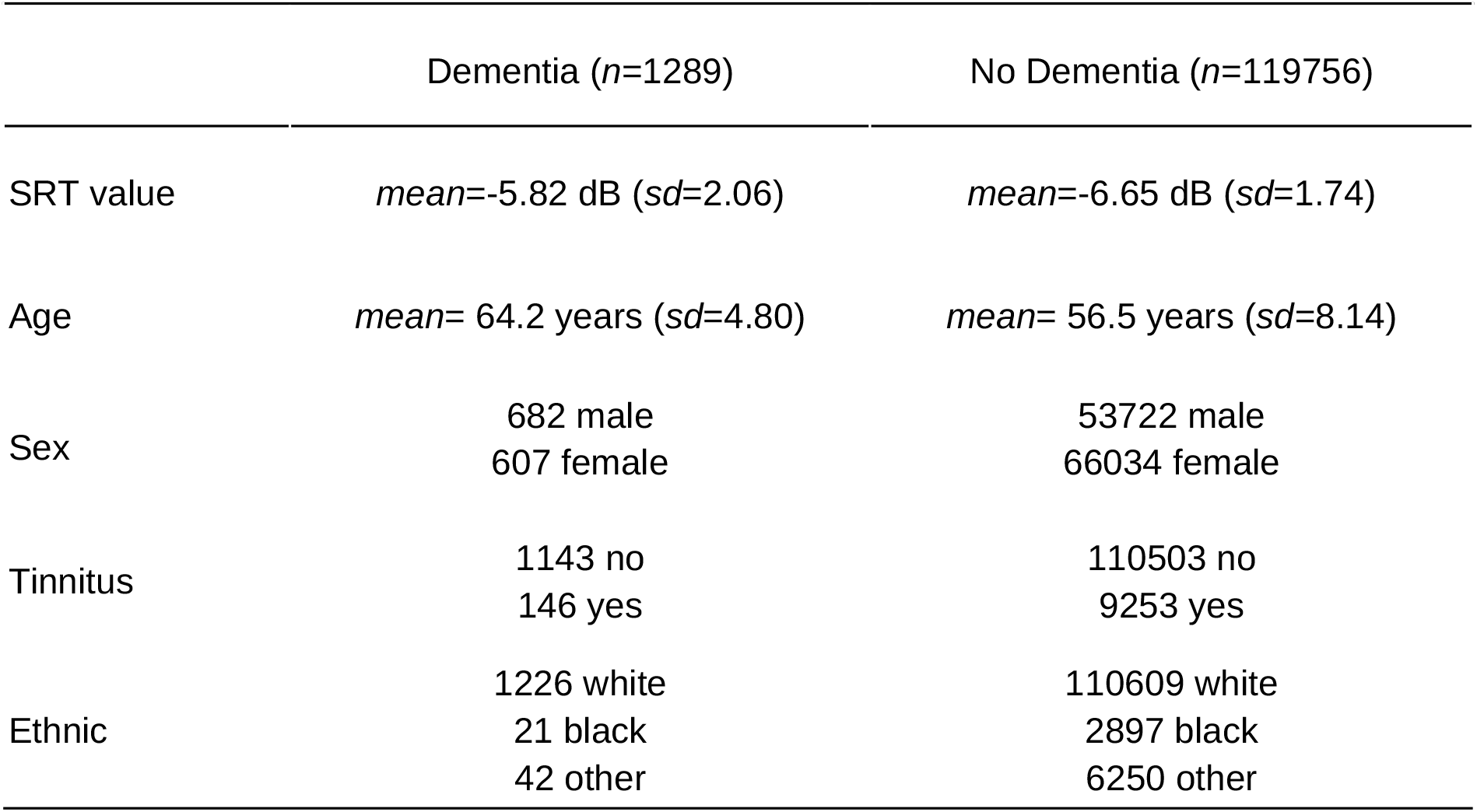
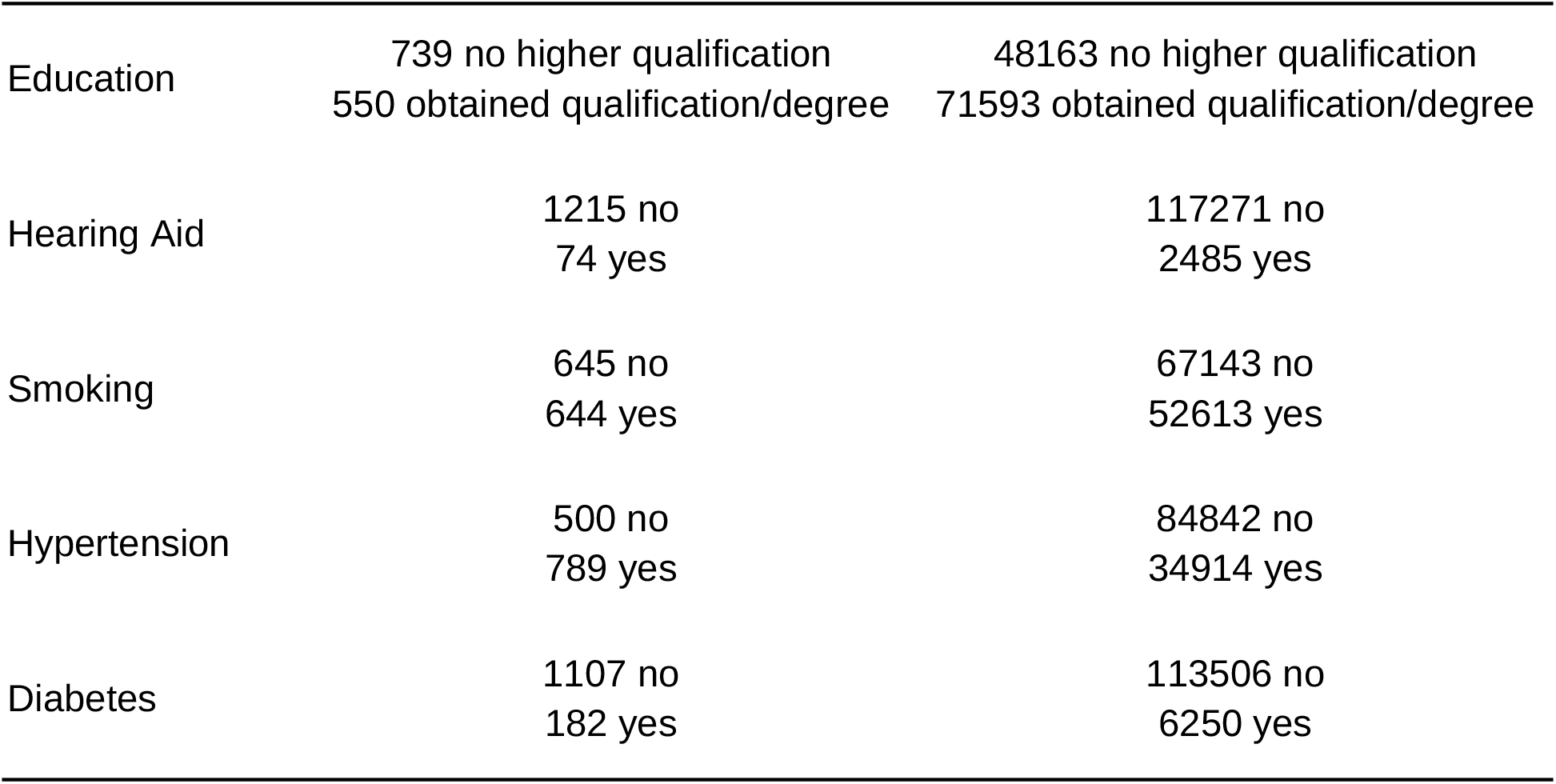

|  | Dementia (n=1289) | No Dementia (n=119756) |
| --- | --- | --- |
| SRT value | mean=-5.82 dB (sd=2.06) | mean=-6.65 dB (sd=1.74) |
| Age | mean= 64.2 years (sd=4.80) | mean= 56.5 years (sd=8.14) |
| Sex | 682 male<br>607 female | 53722 male<br>66034 female |
| Tinnitus | 1143 no<br>146 yes | 110503 no<br>9253 yes |
| Ethnic | 1226 white<br>21 black<br>42 other | 110609 white<br>2897 black<br>6250 other |
| Education | 739 no higher qualification<br>550 obtained qualification/degree | 48163 no higher qualification<br>71593 obtained qualification/degree |
| Hearing Aid | 1215 no<br>74 yes | 117271 no<br>2485 yes |
| Smoking | 645 no<br>644 yes | 67143 no<br>52613 yes |
| Hypertension | 500 no<br>789 yes | 84842 no<br>34914 yes |
| Diabetes | 1107 no<br>182 yes | 113506 no<br>6250 yes |

